# Inference of causal relationships between sleep-related traits and 1,527 phenotypes using genetic data

**DOI:** 10.1101/2020.05.06.20092643

**Authors:** Luis M. García-Marín, Adrián I. Campos, Nicholas G. Martin, Gabriel Cuéllar-Partida, Miguel E. Rentería

## Abstract

**Study Objective:** Sleep is essential for both physical and mental health. There is an increasing interest in understanding how different factors shape individual variation in sleep duration, quality and patterns, or confer risk for sleep disorders. The present study aimed to identify novel causal relationships between sleep-related traits and other phenotypes, using a genetics-driven hypothesis-free approach not requiring longitudinal data.

**Methods:** We used genetic data and the latent causal variable (LCV) method to screen the phenome and infer causal relationships between seven sleep-related traits (insomnia, daytime dozing, easiness of getting up in the morning, snoring, sleep duration, napping, and morningness) and 1,527 different phenotypes.

**Results:** We identify 84 significant causal relationships. Among other findings, poor health of musculoskeletal and connective tissue disorders increase insomnia risk and reduce sleep duration; depression-related traits increase insomnia and daytime dozing; insomnia, napping and snoring are affected by obesity and cardiometabolic traits and diseases; and working with asbestos, thinner, or glues increases insomnia, potentially through an increased risk of respiratory disease.

**Conclusion:** Overall, our results indicate that changes in sleep variables are predominantly the consequence, rather than the cause, of other underlying phenotypes and diseases. These insights could inform the design of future epidemiological and interventional studies in sleep medicine and research.

## Introduction

Sleep is a complex neurological and physiological state that is essential for various biological processes and systems, ranging from homeostasis and restoration of energy levels to memory consolidation.^1^ Individual differences in sleep quality, duration, and patterns have been associated with numerous variables, including sex, age, genetics, body size, occupation, mental and physical illness status, and cultural and environmental factors.^2^ Furthermore, growing evidence suggests that inadequate sleep and sleep disorders contribute to the aetiology of metabolic, psychiatric and neurodegenerative conditions.^3–5^

Epidemiological studies of sleep have focused on a few measurable traits, such as sleep duration, snoring, chronotype, and daytime dozing, and diseases such as insomnia and sleep apnoea.^6,7^ Twin and family studies have shown that genetic factors partly explain individual variation in sleep-related traits, with heritability estimates ranging from 0.30 for insomnia and 0.34 for daytime dozing,^8^ to 0.42 for morningness ^9^ and 0.52 for snoring.^10,11^

More recently, genome-wide association studies (GWAS) have achieved considerable progress in the characterisation of the genetic architecture of sleep-related traits.^12–15^ GWAS have identified 351 genomic risk loci associated with chronotype,^13^ 41 genomic loci associated with snoring,^15^ and 202 genomic loci associated with insomnia.^16^ Furthermore, significant genetic overlap with other conditions has been uncovered. For instance, genetic correlations (r_G_) were found between insomnia and depressive symptoms, major depression, anxiety disorder, higher neuroticism scores and lower subjective well-being scores.^16^ While snoring was genetically correlated with sleep apnoea, excessive daytime sleepiness, body mass index (BMI), daytime dozing, schizophrenia, anorexia nervosa and neuroticism score, among others.^15^

A genetic correlation between two polygenic traits may be due to horizontal pleiotropic effects, and therefore is not necessarily indicative of a causal relationship.^17^ Horizontal pleiotropy poses challenges to statistical methods designed to infer causal relationships between environmental exposures and health outcomes using genetic data, such as Mendelian randomisation (MR). Besides pleiotropy, MR can also become limited by weakly-powered discovery GWAS studies, which increase the likelihood of false-positive findings.^17,18^ Given the limitations of MR, alternative methods such as Latent Causal Variable (LCV) have been developed.^17^ LCV mediates the genetic correlation between two traits through a latent variable that has a causal effect on each trait. The LCV method distinguishes between genetic correlation due to horizontal pleiotropy and full or partial genetic causation by estimating the genetic causality proportion (GCP), a parameter that can range from 0 (no genetic causality) to 1 (full genetic causality of trait A on trait B) or −1 (full genetic causality of trait B on trait A).^17^

Due to the importance of sleep in human health, there is growing interest in understanding the determinants of sleep-related traits and their relationships with other health conditions. Previous studies have reported associations between tobacco smoking, alcohol consumption, and body mass index with an increased risk of snoring;^15^ insomnia and an increased risk of depression, diabetes, and cardiovascular disease;^16^ longer sleep duration with an increased risk of breast cancer in women^19^ and shorter sleep duration with a higher risk for myocardial infarction.^20^ Nonetheless, most of these studies lack the design principles to perform causal inferences. In fact, many interventional studies on sleep would be considered unethical.

In the present study, we sought to explore the potential causal relationships of sleep-related traits with a broad spectrum of variables, using new statistical methods to infer causation which rely single nucleotide polymorphism (SNP) data from well-powered GWAS for pairs of traits measured on the same, or different samples. We leverage the extensive collection (n=1,527) of GWAS summary statistics in the Complex Traits Genetics Virtual Lab (CTG-VL) to conduct a hypothesis-free phenome-wide screening of variables causally associated with seven sleep-related phenotypes: insomnia, daytime dozing, easiness to get up, snoring, sleep duration, napping, and morningness. Our results confirm some of the causal associations hypothesised through observational studies, and provide new insights into the relationships between sleep, lifestyle and health.

## Methods

### Datasets

The present study used summary statistics from genome-wide association studies (GWAS) for the seven sleep phenotypes under investigation. The summary statistics resulting from a genome-wide scan summarise relevant parameters including allele frequency, effect size, standard error and the p-value of each genetic variant tested on the trait of interest. Most published GWAS have made their summary statistics available to the scientific community, which enables researchers to leverage previous findings to advance knowledge in distinct fields. The GWAS summary statistics used here correspond to studies of snoring, insomnia, daytime dozing, getting up, sleep duration, napping, and morningness (information for each study is listed in **Table 1**). Datasets were obtained from the repositories reported in their corresponding publications (Campos and García-Marín *et al*. 2019 and Jansen *et al*. 2018)^15,16^

**Table 1.**
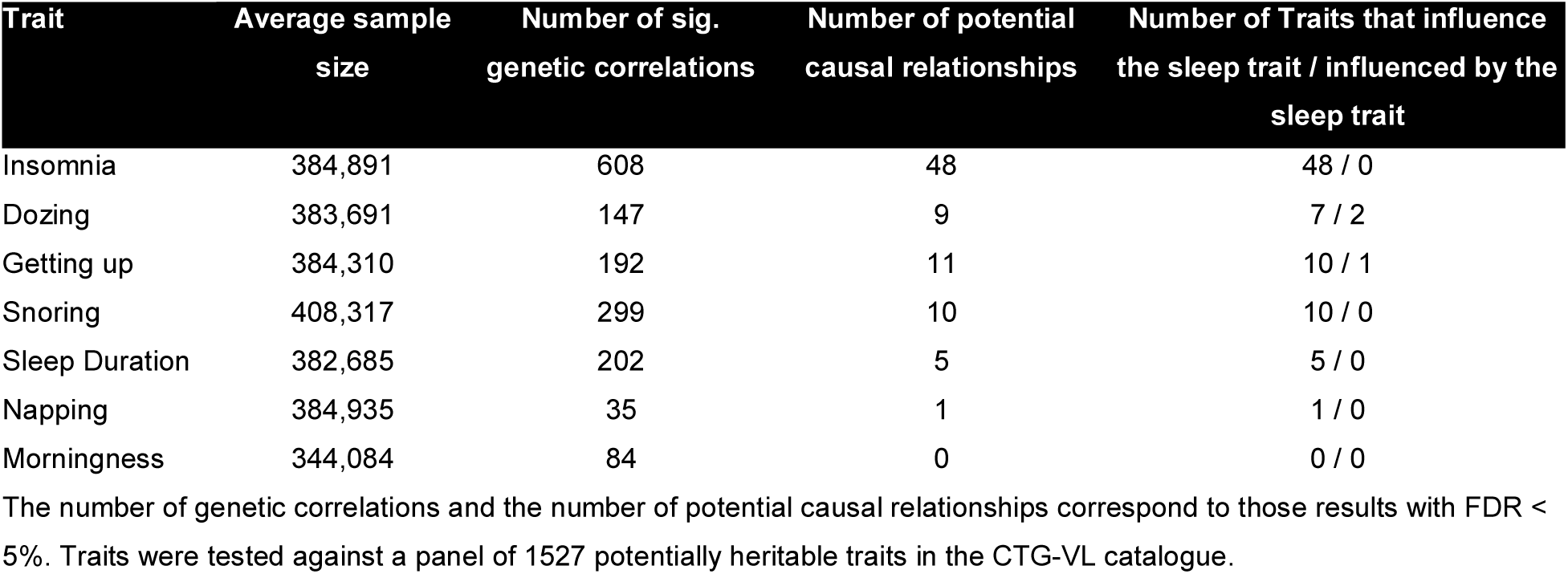
LCV method summary results for each sleep-related trait.

| Trait | Average sample size | Number of sig. genetic correlations | Number of potential causal relationships | Number of Traits that influence the sleep trait / influenced by the sleep trait |
| --- | --- | --- | --- | --- |
| Insomnia | 384,891 | 608 | 48 | 48 / 0 |
| Dozing | 383,691 | 147 | 9 | 7 / 2 |
| Getting up | 384,310 | 192 | 11 | 10 / 1 |
| Snoring | 408,317 | 299 | 10 | 10 / 0 |
| Sleep Duration | 382,685 | 202 | 5 | 5 / 0 |
| Napping | 384,935 | 35 | 1 | 1 / 0 |
| Morningness | 344,084 | 84 | 0 | 0 / 0 |

The number of genetic correlations and the number of potential causal relationships correspond to those results with FDR < 5%. Traits were tested against a panel of 1527 potentially heritable traits in the CTG-VL catalogue.

### Causal architecture analysis pipeline

Summary statistics from genome-wide association studies (GWAS) for seven sleep-related traits were collected from previous studies (Jansen *et al*. 2018 and Campos and García-Marín *et al*. 2019)^15,16^. Then, they were formatted using in-house scripts and uploaded onto the Complex Trait Genomics Virtual Lab (CTG-VL; https://genoma.io/) web-based platform. Subsequently, the *MASSIVE* analysis pipeline, which includes bivariate LD-score regression and latent causal variable analysis (LCV) was implemented for each sleep-related trait of interest. Finally, causal architecture plots were used to depict the LCV results (**Figure 1**).

**Figure 1.**
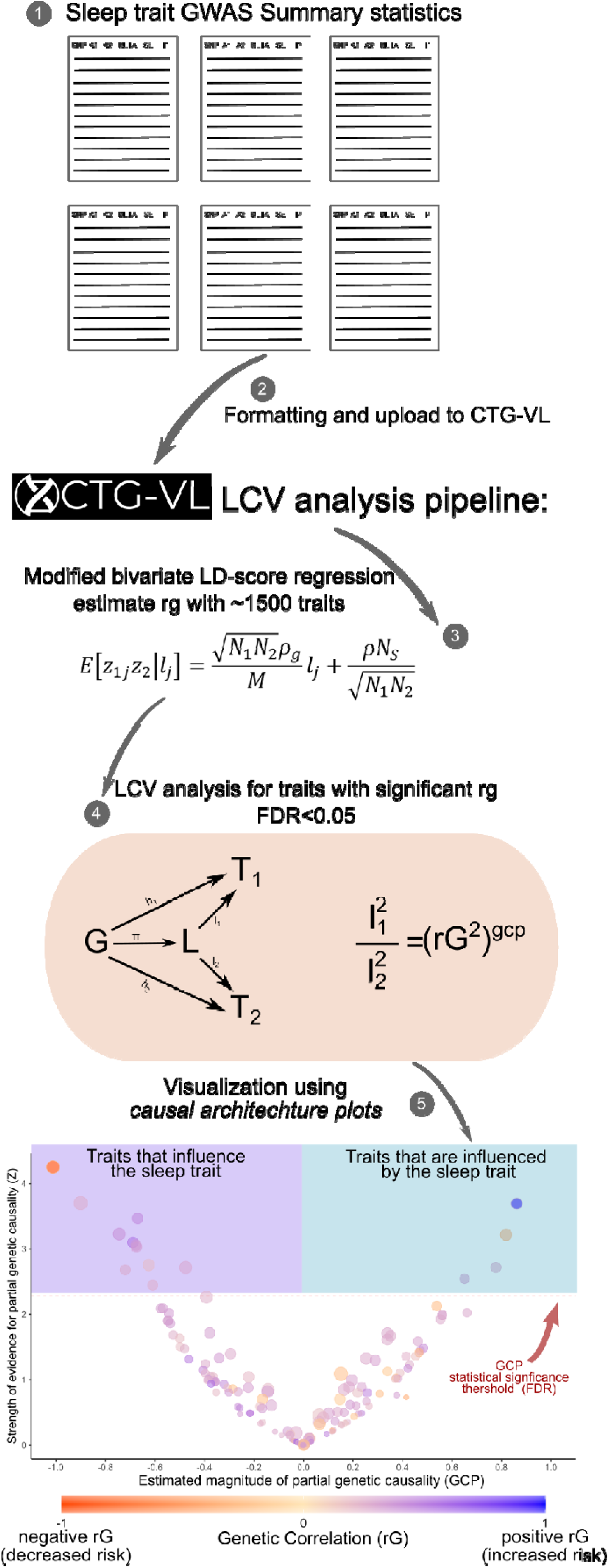
Overview of the causal architecture analysis pipeline. (1) Summary statistics from genome-wide association studies (GWAS) for seven sleep-related traits were obtained from previously published studies. (2) Formatting and loading of data to the Complex Trait Genomics Virtual Lab (CTG-VL). (3) Implementation of the *MASSIVE* analysis pipeline, including bivariate LD-score regression and (4) latent causal variable analysis (LCV). (5) Causal architecture plots were used to describe the LCV results. Each dot represents a trait with a significant genetic correlation. The x-axis shows the estimated genetic causal proportion (GCP) from a latent causal variable. Traits with a positive GCP are influenced by the sleep-related trait studied, whereas traits with a negative GCP are influencing the sleep-related trait of interest. The position on the y-axis represents the strength of the association and a red dashed line shows the statistical significance threshold accounting for multiple testing (FDR < 5%). The LCV diagram of step 4 is based on a diagram from the original LCV publication (O’connor and Price, 2018).

### Genetic correlations

A genetic correlation between two traits describes the relationship of genetic effects sizes at mutual genetic variants across two different phenotypes.^21^ The LCV method estimates a genetic correlation between traits A and B through a modified linkage disequilibrium score regression.^22^ If the genetic correlation is nominally significant, then a latent variable *L* is introduced into the model to assess causality between trait A and trait B, assuming that L is the causal component that mediates the genetic correlation between both traits (see below).^17,18^ We corrected for multiple testing using Benjamini-Hochberg’s False Discovery Rate (FDR < 5%).

### Genetic causal proportion

Latent causal variable (LCV) is a method that uses summary statistics from genome-wide association studies to estimate the genetic causality proportion (GCP) parameter, by mediating the genetic correlation between Trait A and Trait B with a latent variable *L*. A GCP of 0 indicates no genetic causality. A GCP of 1 or −1 is indicative of full genetic causality, of Trait A on Trait B, or Trait B on Trait A, respectively. Values between 0 and 1 or 0 and −1 would indicate partial genetic causality.^17,18^ Although causality is often thought of as a binary characteristic, the idea of partial genetic causality is consistent with both a causal relationship between two complex traits and the notion of distinct genetic components underlying complex traits (e.g. a disease could be caused by both, direct genetic effects, and genetic predisposition to one or more environmental exposures).

An advantage of the LCV method over other methods such as Mendelian randomisation is that it differentiates horizontal from vertical pleiotropy. The model assumes that given a directed effect of trait A on trait B, the effects of genetic variants underlying trait A are expected to have proportional effects on trait B, but not vice versa. Thus, by mediating the genetic correlation between trait A and trait B through the *L* parameter, one can estimate the GCP (see O’Connor & Price^17^ for more details). A GCP value close to 0 suggests that horizontal pleiotropy mediates the genetic correlation between traits A and B, and thus any intervention targeting trait A should not affect trait B.^17^

Notably, the LCV method assumes no bidirectional causality and no confounding by environmental correlates of genotypes. Therefore, care is required when these assumptions are not met.^18^ Moreover, the most attractive features of this method include that it is robust to sample overlap, has higher statistical power than MR and is unconfounded by horizontal pleiotropy.^18^ Multiple testing in GCP was corrected for using Benjamini-Hochberg’s False Discovery Rate (FDR < 5%).

## Results

### Insomnia

We identified genetic correlations between insomnia and 608 traits (FDR < 5%). Forty-eight of these traits showed a causal effect on insomnia risk (negative GCP estimates), and we did not find any significant causal effect of insomnia on another trait (**Table 1**). The traits with the strongest evidence of causal effect on insomnia were *dyspepsia* (r_G_ = 0.34, GCP = −0.97, p-value_GCP_ = 2.37 × 10^−213^), *often worked with materials containing asbestos* (r_G_ = 0.26, GCP = −0.97, p-value_GCP_ = 7.23 × 10^−202^) and *chest pain during physical activity* (r_G_ = 0.57, GCP = −0.96, p-value_GCP_ = 1 33 × 10^−100^; **Table 2**). Consistently, *rarely/never worked with materials containing asbestos* was causally associated with reduced insomnia risk (r_G_ = −0.44, GCP= −0.92, p-value_GCP_ = 2.30 × 10^−33^; **Figure 2 and Supplementary File 1**).

**Table 2.** Traits with a causal effect on insomnia.

| Trait | GCP | GCP se | GCP pval | rG | rG se | rG pval |
| --- | --- | --- | --- | --- | --- | --- |
| Dyspepsia (ICD10) | -0.972 | 0.031 | 2.37E-213 | 0.344 | 0.116 | 3.01E-03 |
| Often worked with materials containing asbestos | -0.968 | 0.031 | 7.23E-202 | 0.266 | 0.1138 | 1.90E-02 |
| Chest pain felt during physical activity | -0.956 | 0.044 | 1.33E-100 | 0.575 | 0.1944 | 3.07E-03 |
| Other disorders of fluid, electrolyte and acid-base balance (ICD10) | -0.910 | 0.076 | 5.17E-33 | 0.378 | 0.1364 | 5.59E-03 |
| Difficulty concentrating during worst period of anxiety | -0.870 | 0.080 | 1.83E-27 | 0.313 | 0.1357 | 2.09E-02 |
| Self-reported type 2 diabetes | -0.934 | 0.095 | 8.66E-23 | 0.233 | 0.100 | 1.98E-02 |
| Self-reported sciatica | -0.701 | 0.090 | 6.63E-15 | 0.412 | 0.179 | 2.16E-02 |
| Why stopped smoking: Illness or ill health | -0.856 | 0.111 | 1.64E-14 | 0.455 | 0.141 | 1.29E-03 |
| Small vessel stroke (Europeans only) | -0.881 | 0.122 | 6.51E-13 | 0.311 | 0.120 | 1.35E-02 |
| Destinations on discharge from hospital: Transfer to other NHS provider: General ward, young physically disabled, A&E | -0.769 | 0.107 | 7.69E-13 | 0.426 | 0.155 | 6.71E-03 |
| Endocrine, nutritional and metabolic diseases | -0.810 | 0.131 | 6.27E-10 | 0.401 | 0.150 | 9.19E-03 |
| Other arthrosis | -0.793 | 0.149 | 1.17E-07 | 0.513 | 0.130 | 1.11E-04 |
| Workplace sometimes very noisy | -0.791 | 0.149 | 1.23E-07 | 0.362 | 0.120 | 2.64E-03 |
| Cylindrical power (left) | -0.768 | 0.147 | 1.95E-07 | 0.224 | 0.097 | 2.17E-02 |
| Fibroblastic disorders | -0.784 | 0.153 | 3.24E-07 | 0.189 | 0.061 | 2.20E-03 |
| Fractured bone site(s): Other bones | -0.758 | 0.153 | 8.41E-07 | 0.264 | 0.081 | 2.68E-03 |
| Self-reported cervical spondylosis | -0.767 | 0.159 | 1.59E-06 | 0.392 | 0.119 | 1.05E-03 |
| Other gastritis (incl. Duodenitis) | -0.767 | 0.162 | 2.35E-06 | 0.506 | 0.097 | 2.20E-07 |
| Fibroblastic disorders (ICD10) | -0.760 | 0.168 | 6.36E-06 | 0.186 | 0.064 | 3.49E-03 |
| Why stopped smoking: Doctor's advice | -0.756 | 0.168 | 7.20E-06 | 0.397 | 0.104 | 1.44E-04 |
| Mouth/teeth dental problems: Toothache | -0.726 | 0.166 | 1.23E-05 | 0.315 | 0.071 | 9.56E-06 |
| Mouth/teeth dental problems: Mouth ulcers | -0.739 | 0.170 | 1.51E-05 | 0.200 | 0.046 | 1.86E-05 |
| Primary gonarthrosis (Bilateral) | -0.733 | 0.171 | 1.93E-05 | 0.298 | 0.117 | 1.11E-02 |
| Treatment/medication: Diazepam | -0.740 | 0.174 | 2.13E-05 | 0.443 | 0.170 | 9.47E-03 |
| Mouth ulcers | -0.723 | 0.170 | 2.24E-05 | 0.193 | 0.042 | 5.49E-06 |
| Knee pain for 3+ months | -0.718 | 0.181 | 7.46E-05 | 0.395 | 0.101 | 1.04E-04 |
| Diseases of the ear and mastoid process | -0.718 | 0.183 | 9.14E-05 | 0.301 | 0.116 | 9.90E-03 |
| Workplace sometimes very dusty | -0.718 | 0.194 | 2.13E-04 | 0.489 | 0.149 | 1.07E-03 |
| Other specific joint derangements/joint disorders | -0.728 | 0.202 | 3.21E-04 | 0.647 | 0.131 | 7.92E-07 |
| Palmar fascial fibromatosis [Dupuytren] | -0.702 | 0.206 | 6.72E-04 | 0.165 | 0.059 | 5.40E-03 |
| Ever heard an un-real voice | -0.677 | 0.203 | 8.63E-04 | 0.265 | 0.106 | 1.26E-02 |
| ILD differential diagnosis | -0.648 | 0.214 | 2.49E-03 | 0.37 | 0.059 | 2.59E-10 |
| COPD differential diagnosis | -0.648 | 0.214 | 2.49E-03 | 0.37 | 0.059 | 2.59E-10 |

This table shows all traits with a significant (FDR < 5%) genetic causal proportion (GCP) for insomnia with a positive genetic correlation ( $r_G > 0$ ) and a strong GCP estimate (GCP < -0.60). Results for all nominally significant genetic correlations for insomnia are reported in **Supplementary File 1**.

**Figure 2.**
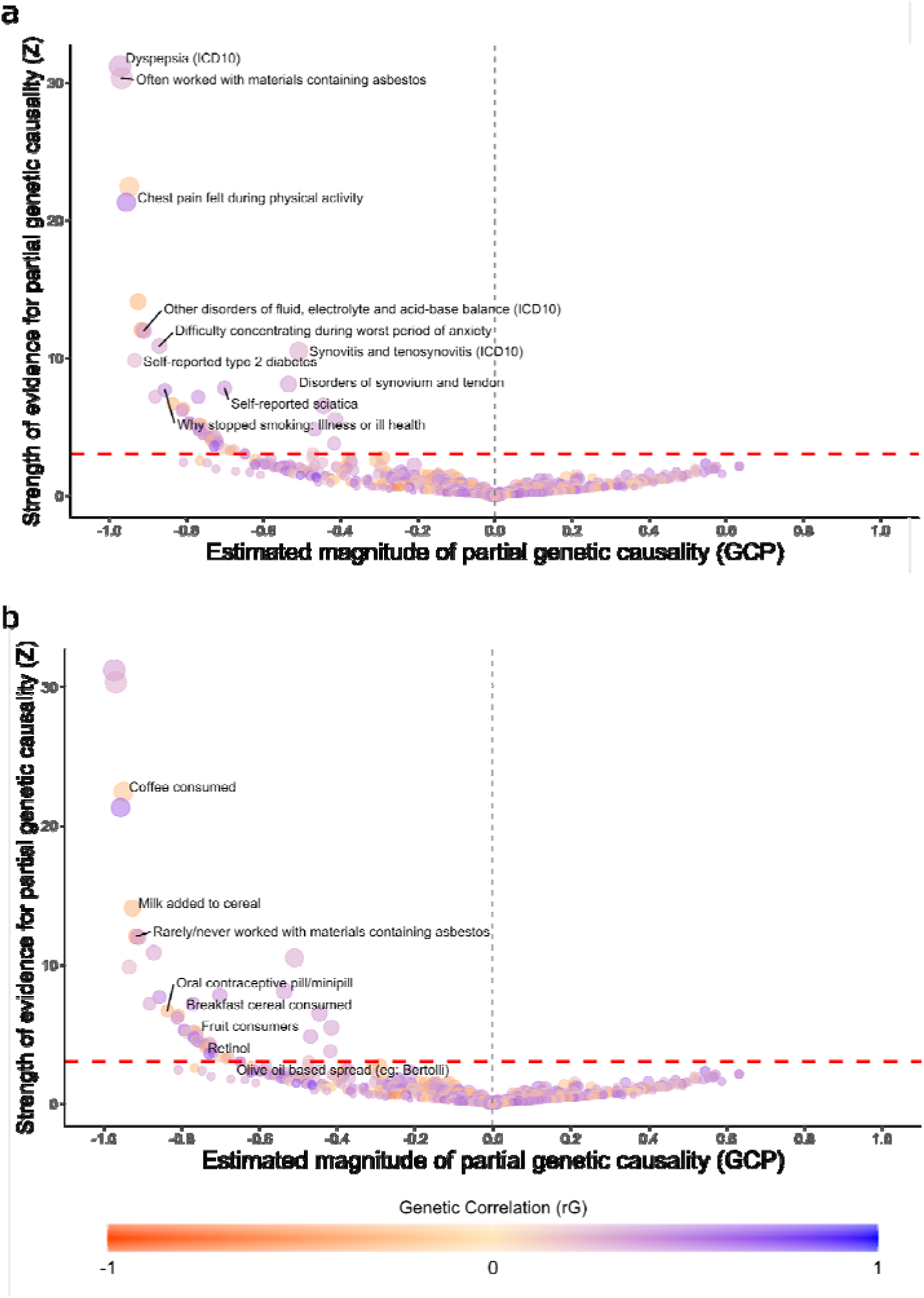
Causal associations for insomnia. Causal architecture plots depicting the latent causal variable exposome-wide analysis results. Dots represent traits with a significant genetic correlation with the trait of interest. The x-axis shows the genetic causality proportion estimate (GCP), while the y-axis shows the GCP absolute Z-score (statistical significance). The red dashed lines represent the statistical significance threshold (FDR<5%) while the grey dashed lines represent the division for traits causally influencing a sleep trait (on the right) and traits causally influenced by the sleep trait (on the left). Showing separately results for traits that increased risk for insomnia (a) and decreased risk for insomnia (b). An explanation for how to interpret these plots is available in **Figure 1**.

Traits related to connective tissue and musculoskeletal health, such as *other specific joint derangements/joint disorders* (r_G_ = 0.64, GCP = −0.73, p-value_GCP_ = 3.21 × 10^−04^), *synovitis* and *tenosynovitis (ICD10)* (r_G_ = 0.28, GCP = −0.50, p-value_GCP_ = 9.65 × 10^−26^) and *other arthrosis* (r_G_ = 0.51, GCP = −0.79, p-value_GCP_ = 1.17 × 10^−07^), were also found to causally influence insomnia risk. Additionally, respiratory-related traits including *interstitial lung disease (ILD)* (r_G_ = 0.38, GCP = −0.65, p-value_GCP_ = 0.003) and *chronic obstructive pulmonary disease (COPD)* (r_G_ = 0.38, GCP = −0.65, p-value_GCP_ = 0.003) showed evidence of increasing the risk for insomnia as did gastrointestinal phenotypes including *other gastritis* (r_G_ = 0.51, GCP = −0.76, p-value_GCP_ = 2.35 × 10^−06^). A similar pattern was observed for *stopped smoking due to an illness* (r_G_ = 0.46, GCP = −0.85, p-value_GCP_ = 1.64 × 10^−14^) *or due to a doctor’s advice* (r_G_ = 0.40, GCP = −0.75, p-value_GCP_ = 7.20 × 10^−06^) (**Figure 2**).

### Daytime dozing

We identified 147 traits with a significant genetic correlation with daytime dozing and 9 with evidence for a causal relationship. Of those, seven influence dozing, and two are putative consequences of it (**Figure 3a**). Moreover, five out of the nine causal relationships directly involve depression, with the trait *no bipolar disorder or depression* showing the most robust evidence for decreasing the risk for daytime dozing (r_G_ = −0.32, GCP = −0.68, p-value_GCP_ = 2.13 × 10^−05^), followed by seeing a doctor (GP) for *nerves, anxiety, tension or depression*, which increased risk for daytime dozing (r_G_ = 0.27, GCP = −0.53, p-value_GCP_ = 2.15 × 10^−04^;**Table 3**).

**Figure 3.**
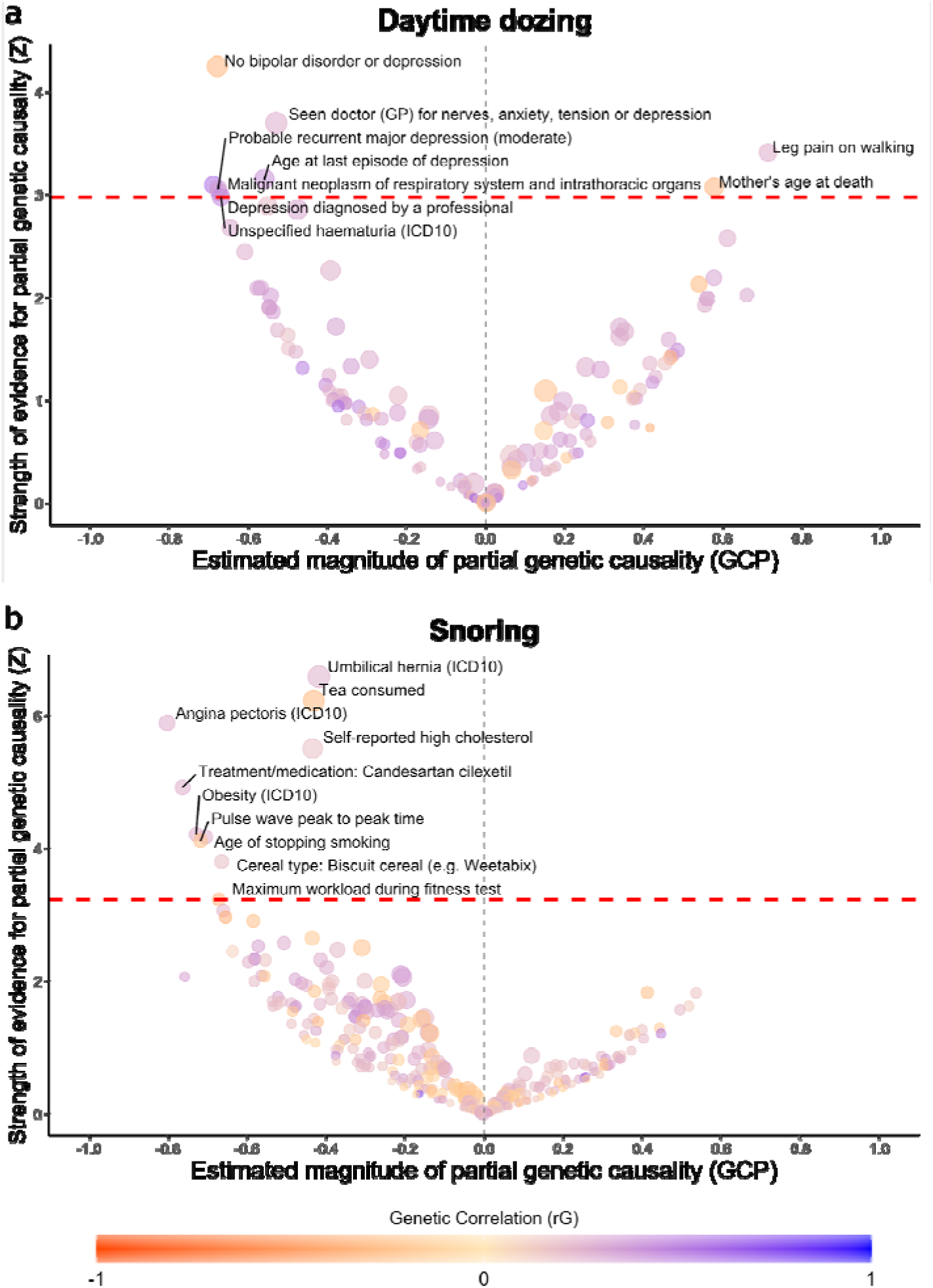
Causal associations for daytime dozing and snoring. Causal architecture plots depicting the latent causal variable exposome-wide analysis results. Dots represent traits with a significant genetic correlation with the trait of interest. The x-axis shows the genetic causality proportion estimate (GCP), while the y-axis shows the GCP absolute Z-score (statistical significance). The red dashed lines represent the statistical significance threshold (FDR<5%) while the grey dashed lines represent the division for traits causally influencing a sleep trait (on the right) and traits causally influenced by the sleep trait (on the left). Showing results for daytime dozing (a) and snoring (b). An explanation for how to interpret these plots is available in **Figure 1**.

**Table 3.** Traits with a causal relationship with sleep-related traits.

| Trait 1 | Trait 2 | GCP | GCP se | GCP pval | rG | rG se | rG pval |
| --- | --- | --- | --- | --- | --- | --- | --- |
| Dozing | No bipolar disorder or depression | -0.678 | 0.159 | 2.13E-05 | -0.323 | 0.089 | 2.79E-04 |
| Dozing | Seen doctor (GP) for nerves, anxiety, tension or depression | -0.530 | 0.143 | 2.15E-04 | 0.274 | 0.060 | 5.80E-06 |
| Dozing | Leg pain on walking | 0.713 | 0.209 | 6.44E-04 | 0.266 | 0.089 | 2.72E-03 |
| Dozing | Age at last episode of depression | -0.559 | 0.177 | 1.59E-03 | 0.403 | 0.141 | 4.25E-03 |
| Dozing | Malignant neoplasm of respiratory system and intrathoracic organs | -0.689 | 0.222 | 1.95E-03 | 0.569 | 0.179 | 1.49E-03 |
| Dozing | Mother's age at death | 0.577 | 0.187 | 2.10E-03 | -0.314 | 0.104 | 2.51E-03 |
| Dozing | Probable recurrent major depression (moderate) | -0.676 | 0.221 | 2.22E-03 | 0.372 | 0.132 | 4.82E-03 |
| Dozing | Depression diagnosed by a professional | -0.674 | 0.222 | 2.46E-03 | 0.345 | 0.082 | 2.25E-05 |
| Dozing | Unspecified haematuria (ICD10) | -0.669 | 0.224 | 2.90E-03 | 0.463 | 0.101 | 4.12E-06 |
| Getting up | Episodal and paroxysmal disorders | -0.823 | 0.052 | 4.64E-55 | -0.215 | 0.071 | 2.30E-03 |
| Getting up | Easily tired during worst period of anxiety | -0.659 | 0.063 | 4.43E-25 | -0.343 | 0.116 | 3.11E-03 |
| Getting up | Manifestations of mania or irritability: Increased creativity | -0.655 | 0.080 | 3.64E-16 | -0.299 | 0.109 | 6.16E-03 |
| Getting up | Tinnitus: Some of the time | -0.816 | 0.132 | 6.38E-10 | -0.354 | 0.107 | 8.94E-04 |
| Getting up | Taken unprescribed medication for depression (more than once) | -0.588 | 0.098 | 2.65E-09 | -0.468 | 0.145 | 1.25E-03 |
| Getting up | Age at first episode of depression | -0.789 | 0.150 | 1.51E-07 | 0.435 | 0.100 | 1.21E-05 |
| Getting up | Self-reported anxiety/panic attacks | -0.273 | 0.062 | 1.22E-05 | -0.311 | 0.077 | 4.95E-05 |
| Getting up | Panic attacks diagnosed by a professional | -0.499 | 0.122 | 4.75E-05 | -0.247 | 0.074 | 7.62E-04 |
| Getting up | Treatment/medication: Citalopram | -0.700 | 0.182 | 1.20E-04 | -0.266 | 0.088 | 2.34E-03 |
| Getting up | Ever had prolonged feelings of sadness or depression | 0.680 | 0.216 | 1.66E-03 | -0.368 | 0.041 | 4.46E-19 |
| Getting up | Been in serious accident believed to be life-threatening | -0.595 | 0.193 | 2.04E-03 | -0.233 | 0.079 | 3.14E-03 |
| Snoring | Umbilical hernia (ICD10) | -0.417 | 0.063 | 4.55E-11 | 0.252 | 0.092 | 5.72E-03 |
| Snoring | Tea consumed | -0.430 | 0.069 | 4.92E-10 | -0.309 | 0.078 | 6.58E-05 |
| Snoring | Angina pectoris (ICD10) | -0.801 | 0.136 | 3.90E-09 | 0.257 | 0.052 | 8.56E-07 |
| Snoring | Self-reported high cholesterol | -0.433 | 0.078 | 3.75E-08 | 0.172 | 0.043 | 6.45E-05 |
| Snoring | Treatment/medication: Candesartan cilexetil | -0.761 | 0.154 | 8.71E-07 | 0.254 | 0.074 | 5.55E-04 |
| Snoring | Obesity (ICD10) | -0.727 | 0.172 | 2.54E-05 | 0.265 | 0.104 | 1.06E-02 |
| Snoring | Age of stopping smoking | -0.704 | 0.168 | 3.06E-05 | 0.270 | 0.098 | 5.75E-03 |
| Snoring | Pulse wave peak to peak time | -0.716 | 0.174 | 3.82E-05 | -0.240 | 0.069 | 5.60E-04 |
| Snoring | Cereal type: Biscuit cereal (e.g. Weetabix) | -0.663 | 0.174 | 1.44E-04 | 0.185 | 0.059 | 1.89E-03 |
| Snoring | Maximum workload during fitness test | -0.670 | 0.207 | 1.24E-03 | -0.364 | 0.081 | 7.43E-06 |
| Sleep duration | Semi skimmed milk consumed | -0.848 | 0.096 | 1.53E-18 | 0.356 | 0.128 | 5.39E-03 |
| Sleep duration | Disorders of synovium and tendon + bursopathies | -0.826 | 0.100 | 2.39E-16 | -0.326 | 0.112 | 3.80E-03 |
| Sleep duration | Treatment/medication: Movicol oral powder | -0.851 | 0.120 | 1.46E-12 | -0.437 | 0.153 | 4.12E-03 |
| Sleep duration | Primary gonarthrosis (Bilateral) | -0.789 | 0.147 | 9.34E-08 | -0.350 | 0.125 | 5.17E-03 |
| Sleep duration | Factors influencing health status and contact with health services | -0.689 | 0.203 | 6.94E-04 | -0.204 | 0.072 | 4.70E-03 |
| Napping | Triglycerides | -0.833 | 0.108 | 1.30E-14 | 0.159 | 0.037 | 1.52E-05 |

This table shows all traits with a significant (FDR < 5%) genetic causal proportion (GCP) for daytime dozing, dozing, ease to get up in the morning, snoring, sleep duration and napping. Nominally significant genetic correlations for all sleep-related traits are reported in **Supplementary File 1**.

### Snoring

S*noring* was genetically correlated with 299 different traits, ten of which held a causal effect on snoring (**Figure 3b**). The identified causal relationships that increased the risk of snoring included *umbilical hernia* (r_G_ = 0.25, GCP = −0.42, p-value_GCP_ = 4.55 × 10^−11^), *angina pectoris* (r_G_ = 0.26, GCP = −0.80, p-value_GCP_ = 3.90 × 10^−09^) and *obesity* (r_G_ = 0.27, GCP = −0.73, p-value_GCP_ = 2.54 × 10^−05^; **Table 3**), all of which were ascertained as an International Classification of Diseases (ICD10) diagnosis (see discussion).

### Sleep Duration

Two hundred and two traits were genetically correlated with sleep duration. Five of them were found to causally influence sleep duration. Similar to insomnia, traits related to connective tissue and musculoskeletal health showed evidence of a causal association, including *disorders of synovium and tendon + bursophaties* (r_G_ = −0.33, GCP = −0.83, p-value_GCP_ = 2.39 × 10^−16^) and *primary gonarthrosis (bilateral)* (r_G_ = −0.35, GCP = −0.79, p-value_GCP_ = 9.34 × 10^−08^), both decreasing sleep duration (**Figure 4a** and **Table 3)**.

**Figure 4.**
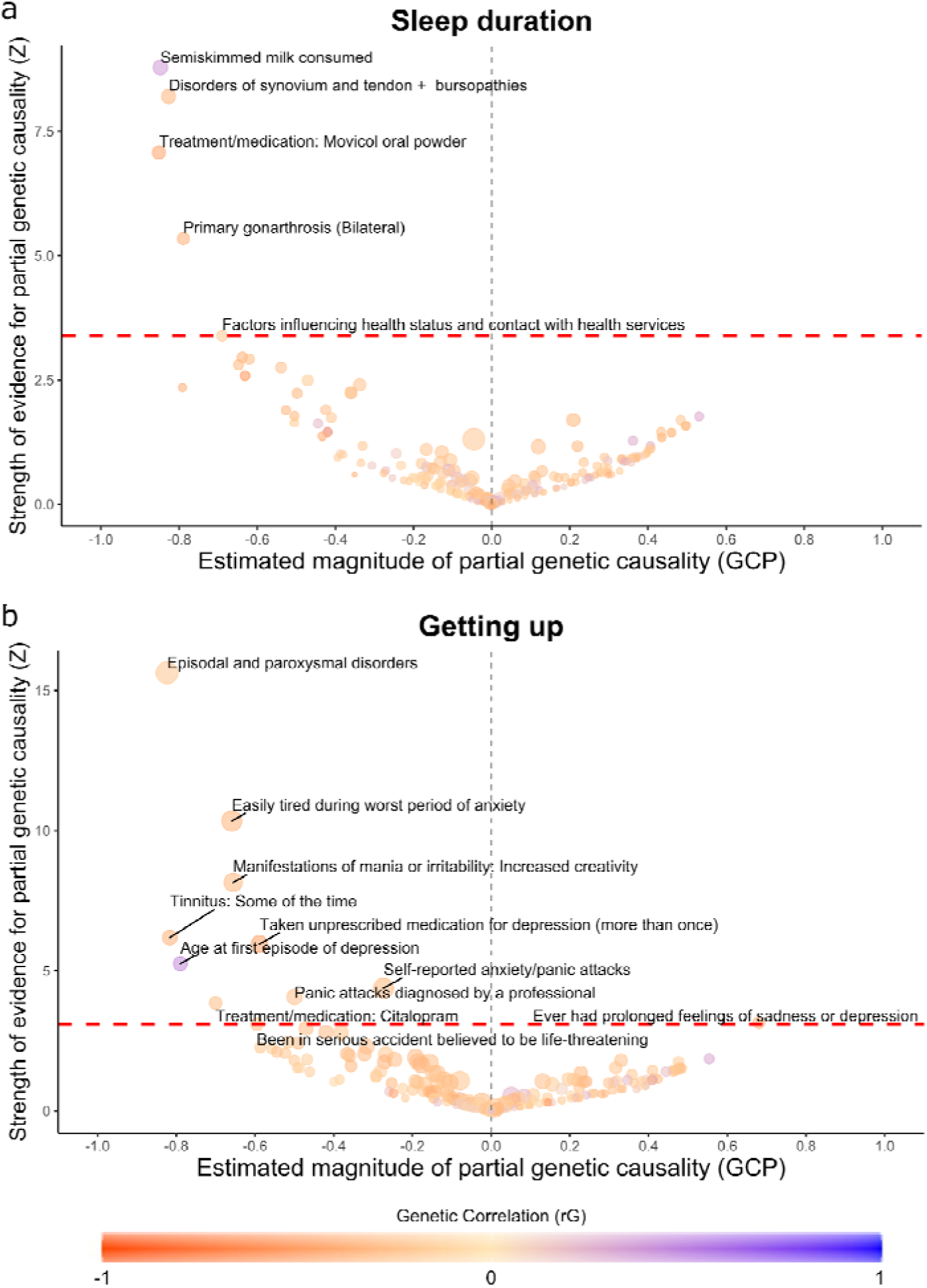
Causal associations for sleep duration and getting up. Causal architecture plots depicting the latent causal variable exposome-wide analysis results. Dots represent traits with a significant genetic correlation with the trait of interest. The x-axis shows the genetic causality proportion estimate (GCP), while the y-axis shows the GCP absolute Z-score (statistical significance). The red dashed lines represent the statistical significance threshold (FDR<5%) while the grey dashed lines represent the division for traits causally influencing a sleep trait (on the right) and traits causally influenced by the sleep trait (on the left). Showing results for sleep duration (a) and ease of getting up (b). An explanation for how to interpret these plots is available in **Figure 1**.

### Getting Up

The *ease of getting up in the morning* was genetically correlated with 192 traits. We found evidence for ten traits causally influencing getting up and one being causally influenced by it. The *age at the first episode of depression* was the only trait associated with being easier for an individual to get up in the morning (r_G_ = 0.44, GCP = −0.79, p-value_GCP_ = 1.51 × 10^−07^). In contrast, the only trait that was influenced by getting up was *ever had prolonged feelings of sadness or depression* (r_G_ = −0.37, GCP = 0.68, p-value_GCP_ = 0.002). Out of the 11 causal relationships that were identified, six of them directly involve either depression, anxiety or panic attacks (**Figure 4b** and **Table 3**).

### Napping and morningness

Out of the 35 traits genetically correlated with napping, only *triglyceride levels* held a significant causal effect that increased the risk for napping (r_G_ = 0.16, GCP = −0.83, p-value_GCP_ = 1.30 × 10^−14^; **Table 3**; **Supplementary Figure 1a**). For morningness, genetic correlations with 84 traits were identified. However, none of them supported a potential causal relationship independent of pleiotropy (**Supplementary Figure 1b** and **Supplementary File 1**).

## Discussion

This study provides new insights into the determinants and consequences of seven sleep-related traits. We examined potential causal associations between sleep-related phenotypes and 1527 traits and identified 84 significant causal relationships based on genetic evidence. Overall, our results indicate that changes in sleep variables are predominantly the consequence, rather than the cause, of other underlying phenotypes and diseases.

We identified causal genetic influences of several conditions on insomnia risk. Consistent with previous studies, ^23,24^ gastrointestinal disorders such as *dyspepsia* and *other gastritis*, including *duodenitis*, and respiratory diseases, increased the risk of insomnia. The effects of asthma on insomnia have been described before, showing that uncontrolled asthma is a risk factor for insomnia.^25,26^ Additionally, *COPD* and with *ILD*, also showed a causal effect on increased insomnia risk. Exposure to *asbestos, dust and substances containing solvents such as paint, thinners and glues* was also a causal factor for insomnia. Asbestos and solvents are hazardous chemicals that induce an inflammatory response in the respiratory system that may lead to *pulmonary fibrosis,^27^ ILD,^28^ COPD,^29^* and lung cancer.^30,31^ Therefore, our results suggest that exposure to asbestos and solvents could lead to insomnia as a consequence of the development of severe respiratory diseases such as *COPD, ILD* and *asthma*.

Musculoskeletal conditions and connective tissue disorders also increased insomnia risk. *Synovitis* and *tenosynovitis* (ICD10), *disorders of synovium and tendon*, self-reported *sciatica, fibroblastic disorders* (ICD10), *self-reported cervical spondylosis, primary gonarthrosis (bilateral)* and *knee pain for more than three months*, among others, could be used as a proxy for poor musculoskeletal and connective tissue health. Previous studies have found an association between shorter sleep duration and insomnia with chronic pain.^32^ In the present study, *sciatica, primary gonarthrosis (bilateral)* and *disorders of synovium and tendon* were also causally associated with shorter sleep duration. We speculate that the discomfort and inflammation arising from problems in the musculoskeletal system and connective tissue may reduce sleep duration and, in cases where these disorders prevail for long periods, this relationship may be mediated by chronic pain. However, more research is needed to understand the intricate relationship between pain and sleep.

Depression and anxiety are common among people with insomnia, and previous studies suggest that insomnia may increase the risk of depression and anxiety.^33,34^ According to the Diagnostic and Statistical Manual of Mental Disorders, Fifth Edition (DSM-5), insomnia is considered a secondary symptom of major depression.^35^ Although our results did not show a direct causal relationship between insomnia and anxiety or depression diagnosis, the *frequency* in the use of Diazepam, a common medication for anxiety disorders^36^ and a proxy for anxiety severity, had a causal association with insomnia. Also, *depression diagnosed by a professional* and *age at last episode of depression* among other depression-related traits, showed a causal relationship increasing the risk for daytime dozing. This is expected, as excessive daytime sleepiness is a common symptom of depression.^37,38^

The relationship between insomnia and cardiometabolic diseases has been described before providing an unclear set of conclusions. Most studies suggest that insomnia, particularly in the context of short sleep duration, poses a risk for cardiometabolic diseases, in particular, hypertension ^39–42^ and diabetes mellitus.^40,43^ In contrast, others suggest that insomnia symptoms are not positively associated with hypertension.^44^ In the present study, results uncovered insomnia-increasing causal relationships with cardiometabolic traits including self-reported type 2 diabetes, electrolyte and acid-base balance, as well as endocrine and metabolic diseases, indicating insomnia is most likely a consequence of these diseases. Furthermore, the causal relationships found with *chest pain during physical activity, the use of treatment/medication: GTN 400 micrograms spray*, which is commonly prescribed for hypertension,^45^ and *small vessel stroke*, which is known to be a consequence of hypertension,^46^ suggest that the development of cardiometabolic disease may be causal for insomnia.

*Mouth ulcers* and *stopped smoking due to an illness or a doctor recommendation* shown insomnia-increasing causal relationships. Nonetheless, these relationships are likely explained by a single causal effect on insomnia. Smoking cessation is accompanied by an abstinence phase, which is well-known as a risk factor for mouth ulcers ^47,48^ and insomnia.^48,49^ We speculate that the apparent causal relationship of mouth ulcers on insomnia is mediated through smoking cessation. Formally testing this hypothesis was outside the scope of the present study.

A negative association between the ease of getting up in the morning and depression has been reported previously.^50^ Our results consistently showed causal relationships for anxiety traits such as *panic attacks diagnosed by a professional* and *use of citalopram*, a common antidepressant,^51,52^ increasing the difficulty to get out of bed in the morning. Further, the *age at the first episode of depression*, which is a proxy for the severity and recurrence of depression,^53^ showed a causal relationship with getting up, where higher age increases the ease of getting up. Consistently, we identified a causal relationship where the risk of *ever having prolonged feelings of sadness or depression* was lower for individuals who can effortlessly get up in the morning. These results agree with the fact that sleep problems and reduced energy are part of the diagnostic criteria for clinical depression.

Relationships between snoring and cardiometabolic traits have been reported before.^15,54^ We previously reported a causal link between BMI and snoring^15^ and putative causal links of whole-body fat mass on snoring risk, and snoring increasing blood pressure and pulse rate.^15^ Other studies had also suggested that snoring is a risk factor for cardiometabolic traits such as hypertension^55^ and angina pectoris.^56^ In this study, we identified several factors that influence snoring risk, including *obesity* (ICD10), *angina pectoris* (ICD10), a known risk factor for coronary heart disease (CHD),^57,58^ *self-reported high cholesterol* and *use of candesartan*, a common drug for treating hypertension.^59^ Although this suggests that coronary heart disease exerts a causal relationship on snoring, we cannot currently rule out whether this is mediated through the genetic component for obesity that underlies CHD. The causal relationship found for *triglycerides* causing *napping* is consistent with previous studies in the Chinese population.^60^ However, no evidence was found between *napping* and *CHD* as described in other studies.^60^ We hypothesize that well-powered GWAS would show a relationship with CHD and obesity, all known to correlate with triglyceride levels.

Some limitations of the present study need to be acknowledged. First, our analyses only employed data from individuals of European ancestry. Given that previous studies have highlighted ethnic differences in sleep-related traits,^61–64^ the generalizability of the results may be limited. Also, the GCP estimates are tied to the statistical power of the GWAS, limiting the capacity to identify causal effects for some traits.^64^ In addition, despite the inclusion of more than 1500 traits, other causal associations not tested here may exist. Further, it is crucial to keep in mind the possible biases or designs of the GWAS involved. For instance, our results implicate several medication use GWAS, however, our results suggest these should be interpreted as a proxy for the underlying disease or symptom requiring the medication. Finally, our study highlights the challenge of dealing with non-pleiotropic horizontal associations, where a third trait may moderate the association between two other traits through a shared genetic component. An example is the association of cardiovascular disease-related phenotypes and snoring, which could be mediated through obesity. While we cannot rule out a direct causal association, the most likely explanation is that obesity causes both snoring and cardiovascular disease through a shared genetic component. This limitation is implicit in the bivariate nature of the LCV approach, and future developments on statistical genetics could leverage causal architecture statistical networks to disentangle confounding effects.

In summary, we provide evidence for the causal architecture of sleep-related traits and show that insomnia, daytime dozing, getting up, snoring, sleep duration and napping are mostly consequences of other phenotypes. Our analyses uncovered the role of musculoskeletal and connective tissue disorders in increasing the risk for insomnia and shorter sleep duration. Also, we show the influence of depression on insomnia getting up, and daytime dozing as well as the role of obesity and potentially cardiometabolic traits and diseases, causing an increased risk for insomnia, napping and snoring. We also observed an influence of diet and lifestyle-related variables such as working with asbestos, thinner or glues on respiratory diseases, which in turn increase insomnia risk. Altogether, our results generate testable hypotheses that, if confirmed, could inform the design of novel treatment and intervention strategies to support better sleep quality and overall health.

## Data Availability

Results will be made publicly available via the Complex Trait Genomics Virtual Lab.

https://genoma.io

## Acknowledgments

We thank our colleague Jackson Thorp for his valuable feedback and for proof-reading the manuscript.

## SUPPLEMENTARY FIGURES

**Supplementary Figure 1:**
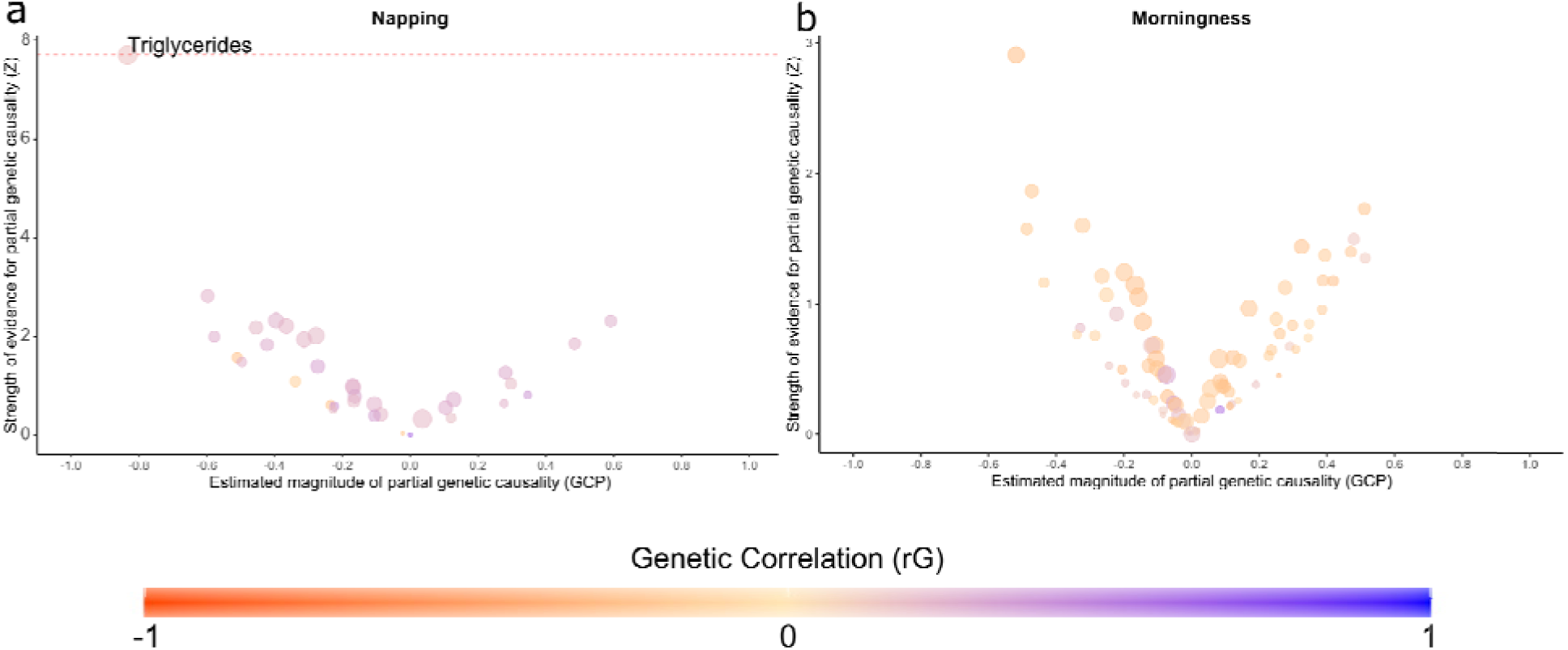
Napping and morningness causal association results. Causal architecture plots depicting the latent causal variable exposome-wide analysis results. Dots represent traits with a significant genetic correlation with the trait of interest. The x-axis shows the genetic causality proportion estimate (GCP), while the y-axis shows the GCP absolute Z-score (statistical significance). The red dashed lines represent the statistical significance threshold (FDR<5%). Showing results for napping (a) and morningness (b). No significant results for morningness. An explanation for how to interpret these plots is available in Figure 1 of the main manuscript.

## SUPPLEMENTARY FILES

**Supplementary File 1**. LCV output for insomnia, dozing, getting up, snoring, sleep duration, napping and morningness.

## REFERENCES

1. Swanson, C. M. et al. The importance of the circadian system & sleep for bone health. Metabolism 84, 28–43 (2018).

2. Troynikov, O., Watson, C. G. & Nawaz, N. Sleep environments and sleep physiology: A review. J. Therm. Biol. 78, 192–203 (2018).

3. Owens, J. A., & Weiss, M. R. Insufficient sleep in adolescents: causes and consequences. Minerva Pediatr. 69, 326–336 (2017).

4. Dong, L., Martinez, A. J., Buysse, D. J. & Harvey, A. G. A composite measure of sleep health predicts concurrent mental and physical health outcomes in adolescents prone to eveningness. Sleep Health 5, 166–174 (2019).

5. Joiner, W. J. The Neurobiological Basis of Sleep and Sleep Disorders. Physiology 33, 317–327 (2018).

6. Ibáñez, V., Silva, J. & Cauli, O. A survey on sleep assessment methods. PeerJ 6, e4849 (2018).

7. Luyster, F. S. et al. Screening and evaluation tools for sleep disorders in older adults. Appl. Nurs. Res. 28, 334–340 (2015).

8. Szily, M. et al. Genetic influences on the onset of obstructive sleep apnoea and daytime sleepiness: a twin study. Respir. Res. 20, 125 (2019).

9. Toomey, R., Panizzon, M. S., Kremen, W. S., Franz, C. E. & Lyons, M. J. A twin-study of genetic contributions to morningness-eveningness and depression. Chronobiol. Int. 32, 303–309 (2015).

10. Watson, N. F., Goldberg, J., Arguelles, L. & Buchwald, D. Genetic and environmental influences on insomnia, daytime sleepiness, and obesity in twins. Sleep 29, 645–649 (2006).

11. Gehrman, P. R. et al. Heritability of insomnia symptoms in youth and their relationship to depression and anxiety. Sleep 34, 1641–1646 (2011).

12. Jones, S. E. et al. Genome-Wide Association Analyses in 128,266 Individuals Identifies New Morningness and Sleep Duration Loci. PLoS Genet. 12, e1006125 (2016).

13. Jones, S. E. et al. Genome-wide association analyses of chronotype in 697,828 individuals provides insights into circadian rhythms. Nat. Commun. 10, 343 (2019).

14. Wang, H. et al. Genome-wide association analysis of self-reported daytime sleepiness identifies 42 loci that suggest biological subtypes. Nat. Commun. 10, 3503 (2019).

15. Campos, A. I. et al. Insights into the aetiology of snoring from observational and genetic investigations in the UK Biobank. Nat. Commun. 11, 817 (2020).

16. Jansen, P. R. et al. Genome-wide analysis of insomnia in 1,331,010 individuals identifies new risk loci and functional pathways. Nat. Genet. 51, 394–403 (2019).

17. O’Connor, L. J., & Price, A. L. Distinguishing genetic correlation from causation across 52 diseases and complex traits. Nat. Genet. 50, 1728–1734 (2018).

18. Haworth, S. et al. Inference and visualization of phenome-wide causal relationships using genetic data: an application to dental caries and periodontitis. bioRxiv 865956 (2020) doi:10.1101/865956.

19. Richmond, R. C. et al. Investigating causal relations between sleep traits and risk of breast cancer in women: mendelian randomisation study. BMJ 365, 12327 (2019).

20. Sleep Duration and Myocardial Infarction. J. Am. Coll. Cardiol. 74, 1304–1314 (2019).

21. Raniti, M. B. et al. Sleep Duration and Sleep Quality: Associations With Depressive Symptoms Across Adolescence. Behav. Sleep Med. 15, 198–215 (2017).

22. Bulik-Sullivan, B. K. et al. LD Score regression distinguishes confounding from polygenicity in genome-wide association studies. Nat. Genet. 47, 291–295 (2015).

23. Lacy, B. E., Everhart, K. & Crowell, M. D. Functional dyspepsia is associated with sleep disorders. Clin. Gastroenterol. Hepatol. 9, 410–414 (2011).

24. Bouchoucha, M. et al. Sleep quality and functional gastrointestinal disorders. A psychological issue. J. Dig. Dis. 19, 84–92 (2018).

25. Sundbom, F., Malinovschi, A., Lindberg, E., Almqvist, C. & Janson, C. Insomnia symptoms and asthma control-Interrelations and importance of comorbidities. Clin. Exp. Allergy 50, 170–177 (2020).

26. Meltzer, L. J., Ullrich, M. & Szefler, S. J. Sleep duration, sleep hygiene, and insomnia in adolescents with asthma. J. Allergy Clin. Immunol. Pract. 2, 562–569 (2014).

27. Gudmundsson, G. & Tomasson, K. {Asbestos and its effects on health of Icelanders - review}. Laeknabladid 105, 327–334 (2019).

28. Gothi, D. et al. Asbestos-induced lung disease in small-scale clutch manufacturing workers. Indian J. Occup. Environ. Med. 20, 95–102 (2016).

29. Wong, J., Magun, B. E. & Wood, L. J. Lung inflammation caused by inhaled toxicants: a review. Int. J. Chron. Obstruct. Pulmon. Dis. 11, 1391–1401 (2016).

30. Konen, T., Johnson, J. E., Lindgren, P. & Williams, A. Cancer incidence and mortality associated with non-occupational and low dose exposure to Libby vermiculite in Minnesota. Environ. Res. 175, 449–456 (2019).

31. Sayan, M. & Mossman, B. T. The NLRP3 inflammasome in pathogenic particle and fibre-associated lung inflammation and diseases. Part. Fibre Toxicol. 13, 51 (2016).

32. Generaal, E., Vogelzangs, N., Penninx, B. W. J. H. & Dekker, J. Insomnia, Sleep Duration, Depressive Symptoms, and the Onset of Chronic Multisite Musculoskeletal Pain. Sleep 40, (2017).

33. Cutler, A. J. The Role of Insomnia in Depression and Anxiety: Its Impact on Functioning, Treatment, and Outcomes. J. Clin. Psychiatry 77, e1010 (2016).

34. Li, L., Wu, C., Gan, Y., Qu, X. & Lu, Z. Insomnia and the risk of depression: a meta-analysis of prospective cohort studies. BMC Psychiatry 16, 375 (2016).

35. Tolentino, J. C. & Schmidt, S. L. DSM-5 Criteria and Depression Severity: Implications for Clinical Practice. Front. Psychiatry 9, 450 (2018).

36. Dhaliwal, J. S., & Saadabadi, A. Diazepam. in StatPearls (StatPearls Publishing, 2020).

37. Hein, M., Lanquart, J.-P., Loas, G., Hubain, P. & Linkowski, P. Prevalence and risk factors of excessive daytime sleepiness in major depression: A study with 703 individuals referred for polysomnography. J. Affect. Disord. 243, 23–32 (2019).

38. Mume, C. O. Excessive daytime sleepiness among depressed patients. Libyan J. Med. 5, (2010).

39. Thomas, S. J., & Calhoun, D. Sleep, insomnia, and hypertension: current findings and future directions. J. Am. Soc. Hypertens. 11, 122–129 (2017).

40. Garg, H. Role of optimum diagnosis and treatment of insomnia in patients with hypertension and diabetes: A review. J Family Med Prim Care 7, 876–883 (2018).

41. Bathgate, C. J. & Fernandez-Mendoza, J. Insomnia, Short Sleep Duration, and High Blood Pressure: Recent Evidence and Future Directions for the Prevention and Management of Hypertension. Curr. Hypertens. Rep. 20, 52 (2018).

42. Javaheri, S. & Redline, S. Insomnia and Risk of Cardiovascular Disease. Chest 152, 435–444 (2017).

43. Depner, C. M., Stothard, E. R. & Wright, K. P., Jr. Metabolic consequences of sleep and circadian disorders. Curr. Diab. Rep. 14, 507 (2014).

44. Vozoris, N. T. Insomnia symptom frequency and hypertension risk: a population-based study. J. Clin. Psychiatry 75, 616–623 (2014).

45. Ghiadoni, L., Magagna, A., Kardasz, I., Taddei, S. & Salvetti, A. Fixed dose combination of perindopril and indapamide improves peripheral vascular function in essential hypertensive patients. Am. J. Hypertens. 22, 506–512 (2009).

46. Prabhakar, P., De, T., Nagaraja, D. & Christopher, R. Angiotensin-converting enzyme gene insertion/deletion polymorphism and small vessel cerebral stroke in Indian population. Int. J. Vasc. Med. 2014, 305309 (2014).

47. Ussher, M., West, R., Steptoe, A. & McEwen, A. Increase in common cold symptoms and mouth ulcers following smoking cessation. Tob. Control 12, 86–88 (2003).

48. Hughes, J. R. Effects of abstinence from tobacco: valid symptoms and time course. Nicotine Tob. Res. 9, 315–327 (2007).

49. Hughes, J. R. & Hatsukami, D. Signs and symptoms of tobacco withdrawal. Arch. Gen. Psychiatry 43, 289–294 (1986).

50. Kripke, D. F. When our body clocks run late: does it make us depressed? Ann Transl Med 4, 178 (2016).

51. Lavretsky, H. et al. Citalopram, methylphenidate, or their combination in geriatric depression: a randomized, double-blind, placebo-controlled trial. Am. J. Psychiatry 172, 561–569 (2015).

52. Sun, Y. et al. Comparative efficacy and acceptability of antidepressant treatment in poststroke depression: a multiple-treatments meta-analysis. BMJ Open 7, e016499 (2017).

53. Pereverseff, R. S., Beshai, S. & Dimova, M. First episode indices associated with lifetime chronicity of depression among formerly depressed participants: an exploratory study. J. Ment. Health 1–7 (2017).

54. Farr, O. M., Rifas-Shiman, S. L., Oken, E., Taveras, E. M. & Mantzoros, C. S. Current child, but not maternal, snoring is bi-directionally related to adiposity and cardiometabolic risk markers: A cross-sectional and a prospective cohort analysis. Metabolism 76, 70–80 (2017).

55. Zou, J. et al. The Relationship between Simple Snoring and Metabolic Syndrome: A Cross-Sectional Study. J Diabetes Res 2019, 9578391 (2019).

56. Endeshaw, Y. et al. Snoring, daytime sleepiness, and incident cardiovascular disease in the health, aging, and body composition study. Sleep 36, 1737–1745 (2013).

57. Carpiuc, K. T., Wingard, D. L., Kritz-Silverstein, D. & Barrett-Connor, E. The association of angina pectoris with heart disease mortality among men and women by diabetes status: the Rancho Bernardo Study. J. Womens. Health 19, 1433–1439 (2010).

58. Celik, A. et al. Presence of angina pectoris is related to extensive coronary artery disease in diabetic patients. Clin. Cardiol. 36, 475–479 (2013).

59. Cho, K. I. et al. Efficacy and Safety of a Fixed-Dose Combination of Candesartan and Rosuvastatin on Blood Pressure and Cholesterol in Patients With Hypertension and Hypercholesterolemia: A Multicenter, Randomized, Double-Blind, Parallel Phase III Clinical Study. Clin. Ther. 41, 1508–1521 (2019).

60. Yang, L. et al. Longer Sleep Duration and Midday Napping Are Associated with a Higher Risk of CHD Incidence in Middle-Aged and Older Chinese: the Dongfeng-Tongji Cohort Study. Sleep 39, 645–652 (2016).

61. Egan, K. J. et al. Amerindian (but not African or European) ancestry is significantly associated with diurnal preference within an admixed Brazilian population. Chronobiol. Int. 34, 269–272 (2017).

62. Chen, X. et al. Racial/Ethnic Differences in Sleep Disturbances: The Multi-Ethnic Study of Atherosclerosis (MESA). Sleep 38, 877–888 (2015).

63. Kaufmann, C. N. et al. Racial/Ethnic Differences in Insomnia Trajectories Among U.S. Older Adults. Am. J. Geriatr. Psychiatry 24, 575–584 (2016).

64. Carnethon, M. R. et al. Disparities in sleep characteristics by race/ethnicity in a population-based sample: Chicago Area Sleep Study. Sleep Med. 18, 50–55 (2016).

